# A Retroactive Study on Factors Influencing the Efficacy of Treatment for Tuberculosis Patients with HIV: based on the data from 2010 to 2020 in Shanghai, China

**DOI:** 10.1101/2023.12.27.23300538

**Authors:** Chenyu Dong, Renfang Zhang, Shenyang Li, Jun Chen, Yunhe Liu, Xiaoqiong Xia, Gang Liu, Yinzhong Shen, Lei Liu, Liyan Zeng

**Author notes:** These authors contributed equally to this work (Chenyu Dong, Renfang Zhang, Shenyang Li). Correspondence: Liyan Zeng, Shanghai Public Health Clinical Center, Fudan University, Intelligent Medicine Institute, Shanghai Medical College, Fudan University, Jinshan District, Shanghai, China, 201508,; Lei Liu Institute of Biomedical Sciences, Shanghai Medical College, Fudan University, Intelligent Medicine Institute, Shanghai Medical College, Fudan University, Shanghai Institute of Stem Cell Research and Clinical Translation, Xuhui District, Shanghai, China, 200032, Yinzhong Shen Shanghai Public Health Clinical Center, Fudan University, Jinshan District, Shanghai, China, 201508.

## Abstract

At present, the factors influencing Tuberculosis (TB) treatment effectiveness in HIV/TB co-infected patients need to be supported by more substantial real-world evidence. A retrospective study is conducted to fill the vacancy. 461 TB patients with HIV are defined as 742 samples according to each TB detection period. 7788 valid treatment records corresponding to 17 drug compositions for TB and 150 clinical indicators with more than 100 records are used to conduct data mining with consensus clustering, Fisher’s exact test, stratified analysis, and three modeling approaches, including logistic regression, support vector machine, and random forest. We find that A CD4^+^ T cell count of 42 cells per μL may serve as a sensitive classification standard for the immune level to assist in evaluating or predicting the efficacy of TB (*P*=0.007); Rifabutin and levofloxacin alone or in combination may be more effective than other first- and second-line anti-TB agents in combination (*P*=0.037); Samples with low immune levels (CD4≦42) may be more resistant to first-line TB drugs (*P*=0.049); Age (*P*=0.015), bicarbonate radical (*P*=0.007), high-density lipoprotein cholesterol (*P*=0.026), pre-treatment CD8^+^ T cell count (*P*=0.015, age<60, male), neutrophil percentage (*P*=0.033, age<60), rifabutin (*P*=0.010, age<60), and cycloserine (*P*=0.027, age<60) may influence the TB treatment effectiveness; More evidence is needed to support the relationship between pre-treatment clinical indicators or drug regimens and TB treatment effectiveness (The best AUC is 0.560∼0.763); The percentage of lymphocytes (*P*=0.028) can be used as an effective TB therapeutic target. These perspectives supplement knowledge in relevant clinical aspects.

## 1 Introduction

TB is the leading cause of death globally from a single infectious agent [1, 2], surpassing HIV. HIV/TB co-infection can accelerate disease progression, whose prevention and treatment become a significant clinical challenge [3–5]. In recent years, updates in prevalence, mortality rate, and treatment guidelines for HIV/TB co-infection mainly focus on drug resistance [6–8]. For patients with drug-sensitive (DS)-TB [9], multidrug-resistant (MDR)-TB [10–13], extensively drug-resistant (XDR)-TB [14], and latent TB, ongoing clinical trials will hopefully transform the landscape for treatment, which are evaluating novel agents, repurposed agents, adjunctive host-directed therapies, and novel treatment strategies [15]. There needs to be more than real-world evidence (RWE) on evaluating TB treatment effects and exploring therapeutic influencing factors in co-infected patients [16]. A small amount of real-world research suggests that TB treatment regimens’ effectiveness is affected by the level of the immune system, based on which more precise TB medical strategies can be provided for co-infected patients with specific immune levels [17, 18].

Achieving the World Health Organization’s End TB Strategy (a 90% decrease in TB incidence and a 95% decrease in TB mortality by 2035 compared with 2015) requires shorter and more effective treatment regimens [19], especially for HIV/TB co-infected individuals [20]. According to the guidelines for diagnosing and treating HIV/AIDS in China, the main anti-tuberculosis drugs are isoniazid, rifampicin, rifabutin, ethambutol, and pyrazinamide. If Mycobacterium tuberculosis is sensitive to first-line anti-tuberculosis agents, a 2-month intensive treatment with “isoniazid + rifampicin (rifabutin) + ethambutol + pyrazinamide” is followed by a 4-month consolidation treatment with “isoniazid + rifampicin (rifabutin)”[21]. The treatment course should be extended to 9 months for patients with delayed response to anti-tuberculosis therapy (the culture of Mycobacterium tuberculosis remains positive after 2 months of treatment) and bone and joint tuberculosis, and to 9 to 12 months for patients with central nervous system tuberculosis (it is recommended to start ART as early as 2 weeks after anti-tuberculosis therapy [22–25]). For co-drug-resistant TB (including MDR-TB or XDR-TB), ART therapy is initiated within 8 weeks of second-line anti-TB drugs. However, there are few retrospective studies on the efficacy evaluation and influencing factors of treatment programs currently [26–28]. The valid conclusions about the factors that may influence the outcome of clinical tests are unclear [29], partly due to the lack of research innovation over the past few decades/to optimize the TB treatment guidelines for HIV patients [30]. Meanwhile, for patients with effective treatment of TB, there is insufficient research evidence for significant recovery of clinical indicators to normal, which means there is still some research space to find potential drug therapeutic targets.

Based on the in-patient and partial out-patient data of HIV and TB comorbidities from 2010 to 2020 in Shanghai Public Health Clinical Center, this study focuses on the immune levels, therapeutic regimens, and other potential clinical indicators that affect the effectiveness of TB treatment for HIV/TB co-infected individuals, providing more real-world research evidence. Multi-factor logistic regression (LR), support vector machine (SVM), and random forest (RF) algorithm are used to construct prediction models for effective TB treatment to supplement the knowledge gap for future personalized treatment. For HIV/TB comorbid patients who have been effectively treated for TB, the preliminary screening of clinical indicators of apparent recovery to normal provides a possible perspective to search for potential drug treatment targets.

## 2 Materials & Methods

### 2.1 Ethics statement

This study is approved by the Ethics Committee of Shanghai Public Clinical Center (No. 2020-S110-01), and the need for signed consent for participation is waived.

### 2.2 Standardized Data

#### 2.2.1 Sample Information

(1) **Sample source**: Data are exported from Shanghai Public Health Clinical Center’s Hospital Information System (HIS), Electronic Medical Record System (EMR), and Laboratory Information System (LIS) and are organized into the original EXCEL table.
(2) **Data cleaning**: Clean, split, integrate, transform, and simplify raw data.
(3) **Enrolled patients**: 461 TB patients with HIV, whose electronic medical records include two or more microbial tuberculosis testing records carried out at different time points and the medication records during the testing period.
(4) **Enrolled samples**: The interval between two consecutive microbial tuberculosis detections in the enrolled patients is regarded as an independent sample, with a total of 742 samples. The statistical results show that the average interval of the included samples is 74.6 days, and the interval that is less than or equal to the average accounts for 80.5%, while 96.64% of the interval is less than or equal to 365 days. Details on the length distribution of the Mtb measurement interval are shown in **Appendix Table S1**.
(5) **Basic information of the samples**: The basic information statistics of the enrolled samples are shown in **Table 1**. Age and gender are uncontrollable factors considered confounding variables in this study, and their interference with the results should be excluded.

#### 2.2.2 Medication Information

(1) **Chemical composition of anti-tuberculosis drugs**: The original drug record is sorted out, and the trade name of the anti-tuberculosis drugs used is replaced with its core chemical composition, as shown in **Figure 1**.
(2) **Drug records statistics and classification principles of the enrolled samples**: According to clinicians’ instructions, 17 drugs for TB are classified into six categories, including rifamycin antibiotics, isoniazid, ethambutol, pyrazinamide, fluoroquinolone, and other second-line anti-TB drugs, as shown for different colors in **Figure 1**. As for the treatment record, there are 4755 (61%) records with both start and end time of medication; the average duration of medication is 11.75 days, and the period of drugs ≧seven days account for 58.3%. Meanwhile, there are 3033 (39%) records with only the start time of medication.

#### 2.2.3 Clinical Information

##### Clinical indicators’ information

After data cleaning and sorting, Clinical features included demographics and routine clinical tests, which are divided into eight categories, including demographics, blood routine examination, urine routine examination, blood biochemical examination, cerebrospinal fluid (CSF) examination, nucleic acid test, immunologic test, routine microbiological examination, as shown in **Table 2**. 150 clinical indicators (with demographics) with more than 100 records are selected for the subsequent step study. In this study, the test results of clinical indicators are discretized. The results within the reference range are denoted as 0 (normal), while those outside the reference range are denoted as 1 (abnormal). Detailed clinical indicators and reference range information are shown in **Figure S1**.

**Figure 1.**
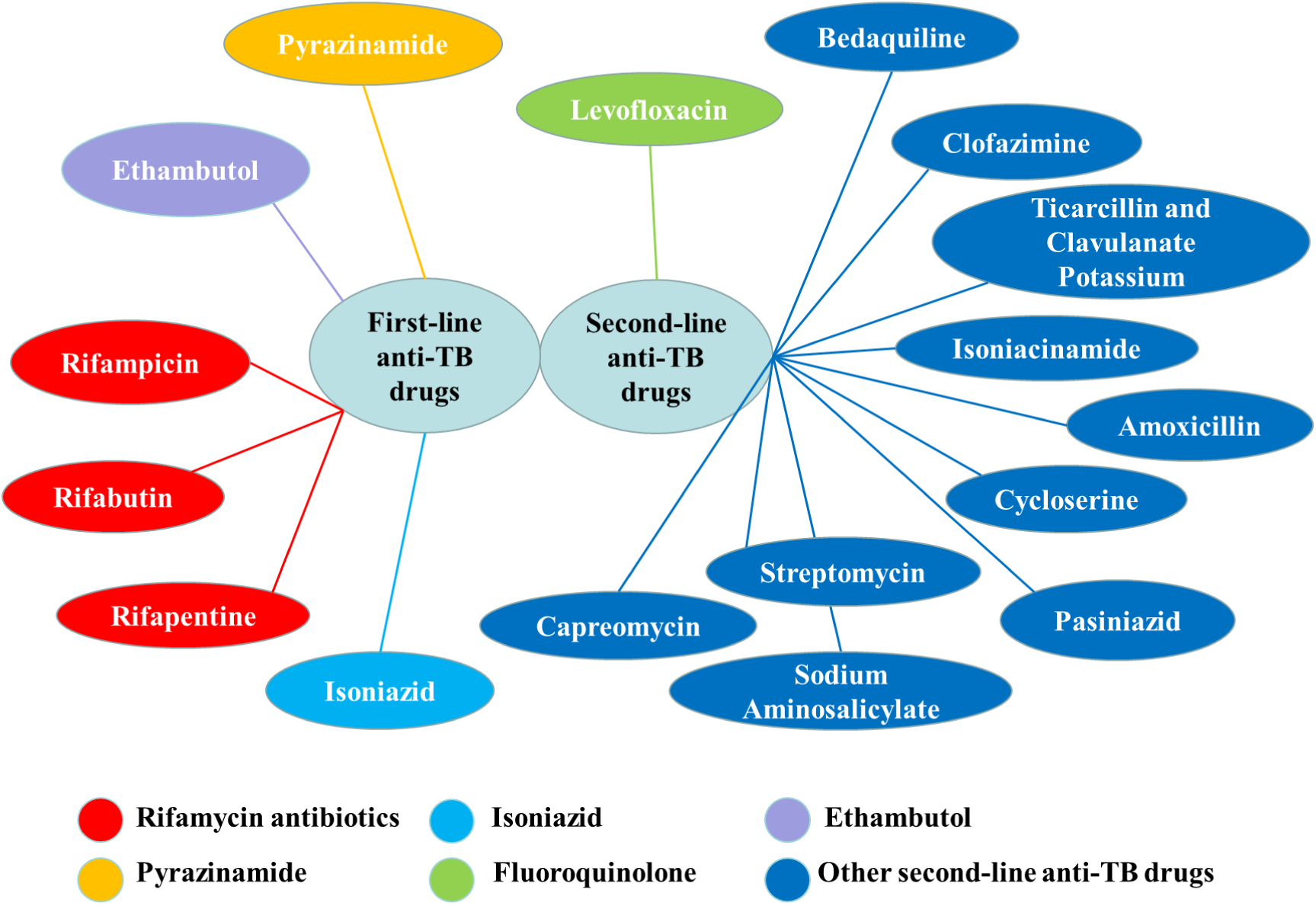
Summary of chemical constituents of anti-tuberculosis drugs.

**Table 1.** Summary of basic information on included samples.

| Basic information | Category | Number | Binary variable |
| --- | --- | --- | --- |
| Age | < 60 | 692 | 0 |
|  | ≥60 | 50 | 1 |
| Gender | male | 682 | 0 |
|  | female | 60 | 1 |
| HBV | no | 739 | 0 |
|  | yes | 3 | 1 |
| Smoking | no | 670 | 0 |
|  | yes | 72 | 1 |
| Drinking | no | 686 | 0 |
|  | yes | 56 | 1 |
| Drug taking | no | 739 | 0 |
|  | yes | 33 | 1 |

**Table 2.** Information summary of clinical indicators category.

| Valid Indicators Category | Number (>100 records) |
| --- | --- |
| Demographics | 6 |
| Blood routine examination | 20 |
| Urine routine examination | 11 |
| Blood biochemical examination | 76 |
| Cerebrospinal fluid (CSF) examination | 22 |
| Nucleic acid test | 4 |
| Immunologic test | 4 |
| Routine microbiological examination | 7 |
| **Total** | **150** |

### 2.3 Retrospective Research Program Design

#### 2.2.1 Screening model of effective sample information

This study designs a set of methods for defining valid samples and screening sample information. Different from the conventional study in which one patient corresponds to one sample unit, in this study, the interval between each two microbial tuberculosis detection time points of a patient is defined as one sample. Therefore, a patient can be considered one or more samples depending on the number of segments. **Figure 2** shows the selection criteria for valid samples and relevant statistical information.

**Figure 2.**
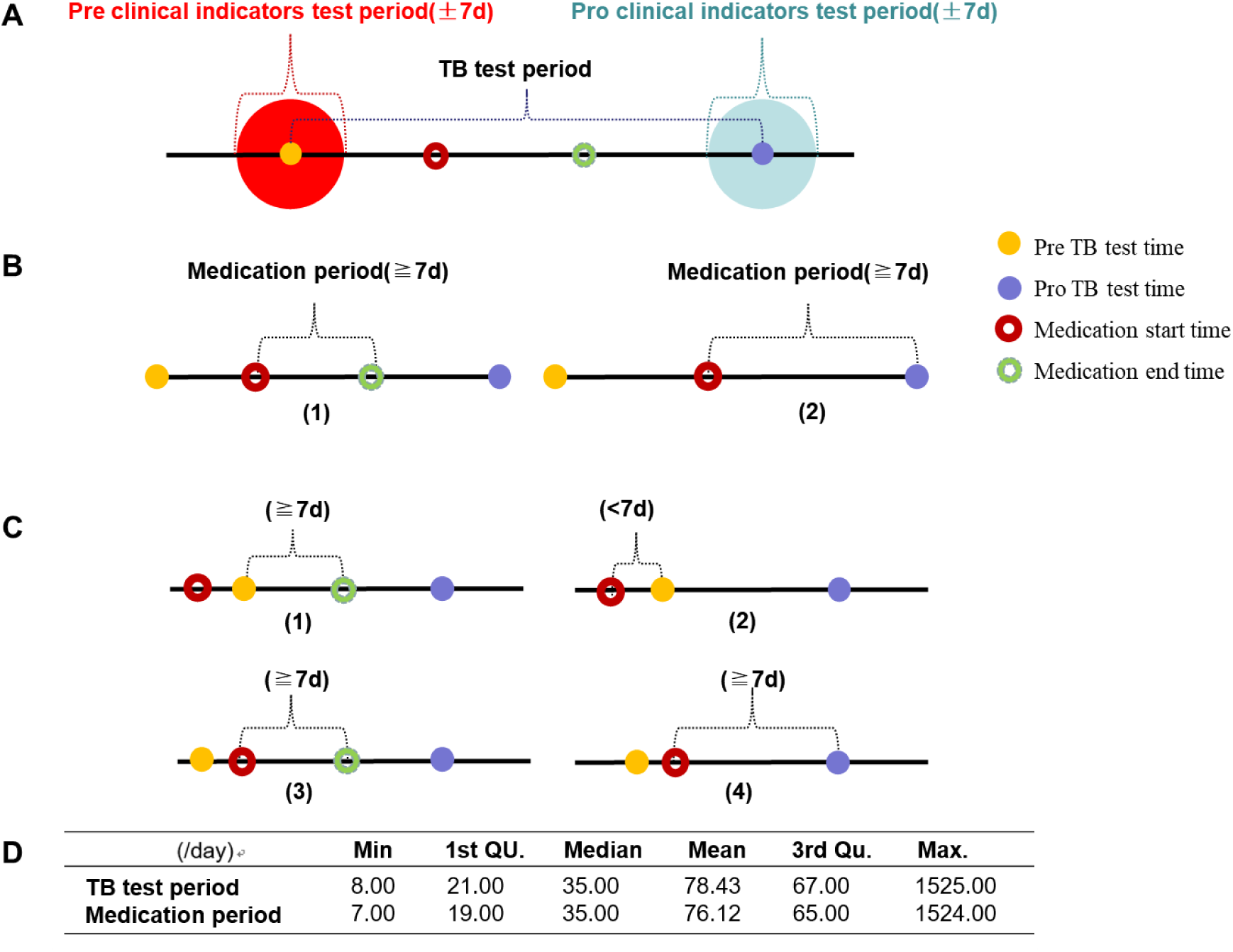
Model of effective sample screening and statistical information.

The screening condition of each clinical indicator is that the indicator test results are within seven days before and after the microbial test time point (values within the reference range are denoted as 0-normal, otherwise denoted as 1-abnormal). If one test record is abnormal, the clinical indicator is denoted as abnormal at that time (**Figure 2A**). According to the relative position of the medication start time and the previous microbial test time, as well as the existence of medication end-time records, screen the valid medication records in each sample unit. For each patient’s first TB test period, the medication start time is the same or later than the first TB test time (**Figure 2B**). For the sample units of patients with more than one TB test period, corresponding to TB detection segments except the first one, the medication start time could be before or later than the pre-TB test time (**Figure 2C**). Drug records that last seven days or more during the TB test period are included in the therapeutic regimen of each sample unit. When calculating the final duration of medication for the same drug that is repeatedly included, if there is only one drug record without end-time, the duration of medication for all the included records of the same drug shall be added together; if there is more than one drug record without end-time, the most prolonged duration among all the drug records without end-time shall be included, followed by adding it to the duration of other drug records with an end-time (**Figure 2B, 2C**). The information of interval duration and corresponding drug duration of samples including drugs are analyzed after the above drug screening of the samples, which conforms to the standards of routine examination time and drug treatment time of TB patients in the official guidelines (**Figure 2D**).

Figure 2A shows the medication and clinical testing information in each sample unit defined in this study. “Pre/Pro TB test time” represents the time point of microbial TB detection before and after treatment. The “Pre/Pro clinical indicators test period(±7d)” represents each clinical indicator’s detection period before and after treatment. Figure 2B shows the screening conditions of drug use records in sample units corresponding to each patient’s first TB test period. Figure 2C shows the drug record screening conditions of the sample units corresponding to other TB detection segments except the first one for patients with more than one TB test period. Figure 2D shows the statistical information of interval duration and corresponding drug duration of 568 samples including drugs after the above drug screening of 742 samples.

#### 2.2.2 Relevant Clinical Trial Criteria

(1) **Criteria for TB disease**: One of the test results of microbial tuberculosis in any sample type and test type is positive[31], not just the sputum specimen.
(2) **Criteria for effective anti-TB drug treatment**: The change of positive to negative microbial tuberculosis test results for one week (≧seven days) or more after taking anti-TB drugs is recorded as 1 (effective drug treatment). In contrast, the persistent positive results are recorded as 0(ineffective drug treatment). Other cases are not included in the analysis.
(3) **Criteria for evaluation of HIV status and efficacy**: CD4^+^ T cell count is considered a key indicator to judge the immune function of HIV-infected persons, and the level of CD4^+^ T cell count in normal people is more than 500 cells / μL. Recent studies have shown that the recovery of the CD4^+^/CD8^+^ ratio to the normal level is expected to be used as a new indicator to evaluate the immune function reconstruction in HIV-infected people[32], and the range of CD4^+^/CD8^+^ in healthy people is 1.5∼2.5. After HIV infection, the number of CD4^+^ T cells decreases significantly while the number of CD8^+^ T cells increases. When the ratio is < 0.5 or < 0.3, the incidence and mortality of HIV-infected persons increase significantly.

#### 2.2.3 Research methods and process

In this study, R 4.1.3 statistical software is used for data processing. Firstly, Fisher’s exact test and stratified analysis are used to test the differences in treatment effectiveness among different medication regimens or groups with different immune statuses. Then, the efficacy prediction models are constructed respectively using logistic regression (LR), support vector machine (SVM), and random forest (RF) algorithm to explore the potential clinical features affecting the therapeutic effect. Model performance is comprehensively evaluated by area under the receiver operating characteristic curve (AUC), accuracy, precision, recall, and F1 score with fivefold cross-validation. When the test set accuracy is the highest, the corresponding data group and model are selected to draw the ROC curve, and the corresponding optimal AUC is displayed. For other evaluation indicators, the mean ± standard deviation results are reported by randomly splitting the training and validation sets five times. Finally, potential biomarkers or therapeutic targets of HIV/TB comorbid patients are explored by comparing the recovery condition of clinical features between the effective treatment group and the ineffective treatment group. All difference analysis results are obtained after controlling for confounding variables of gender and age. The analysis process is shown in Figure 3.

**Figure 3.**
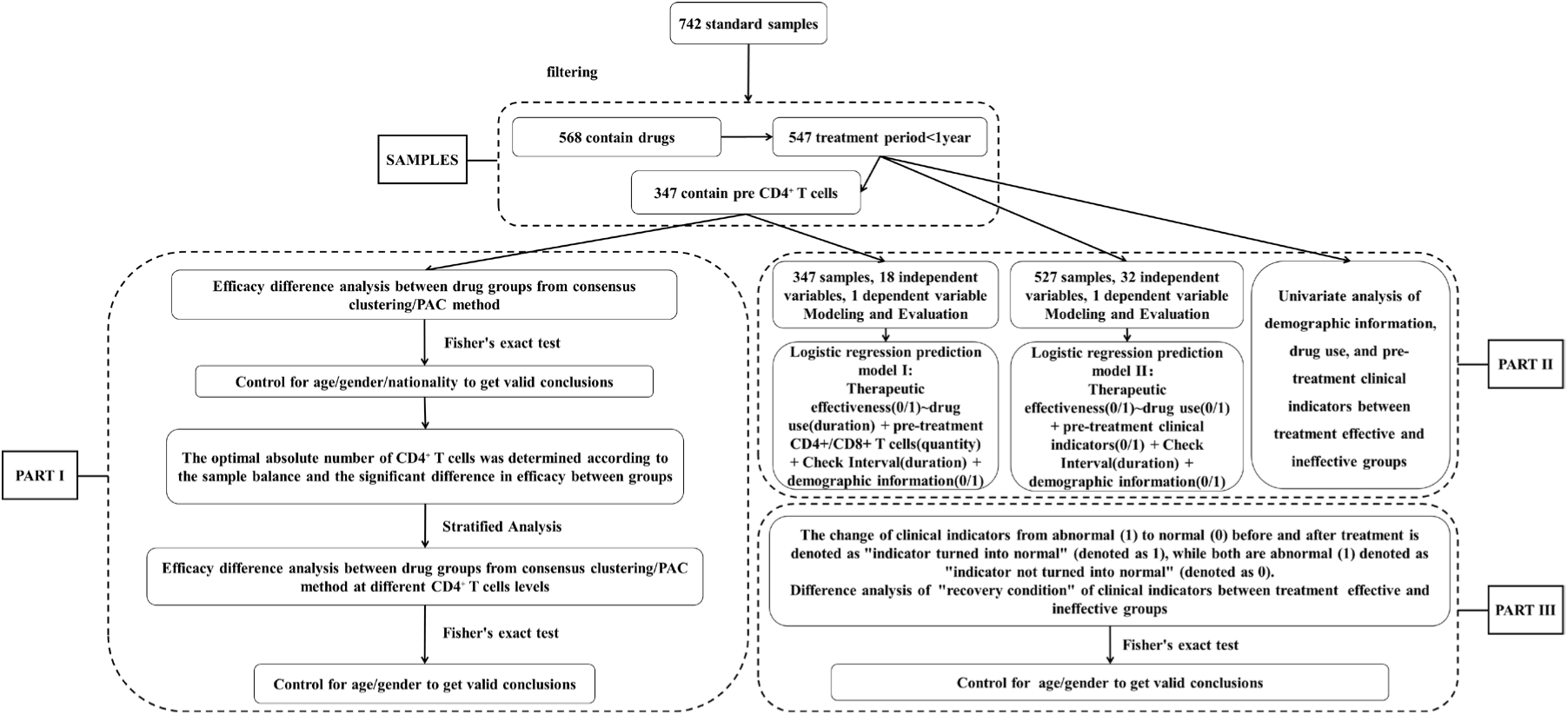
Architecture of the analysis process of the project data.

Figure 3 shows sample filtering and three analysis blocks for this study. Above all, 742 TB standard samples are filtered based on the screening conditions and methods shown in Figure 2, and 568 samples actually containing drug records are obtained. 547 samples with detection intervals of less than one year among the 568 samples are selected as primary valid samples for subsequent research on specific issues. **PART I** shows the steps of comprehensively discussing patients’ immune levels and medication regimens’ influence on TB treatment effectiveness using single-factor hypothesis testing and stratified analysis. **PART II** shows the details of constructing the prediction model of “efficacy - clinical indicators” based on single-factor hypothesis testing and three machine learning algorithms. **PART III** shows the method of using the single-factor hypothesis test to explore initially the clinical indicators which recover from abnormal to normal conditions significantly in the effective TB treatment group compared to the ineffective group.

## 3 Results

### 3.1 Effects of drugs and immune levels on TB treatment effectiveness in HIV/TB co-infected patients

#### 3.1.1 Contribution of CD4^+^ T cell counts

Studies have shown that an individual’s immune level is generally measured by CD4^+^ T cell count[33]. This study first uses the “stepwise method” to determine the best grouping criteria based on CD4^+^ T cell counts. The grouping criteria of CD4^+^ T cell count, the number of samples in each group, and the significant difference in treatment effectiveness between samples in the two groups are shown in **Table S1** (*P*<0.05, Fisher’s exact test).

Considering the size of the *P*-value (the smaller, the better), the balance of the number of samples between the two groups (the more robust, the better) and the interference degree of control variables (the smaller, the better), the CD4^+^ T cell count of 42 cells/uL is finally selected as the optimal grouping criterion to carry out subsequent stratified analysis. When the CD4^+^ T cell count is ≦ 42, the treatment effective rate is 37.71%, and when the CD4^+^ T cell count is > 42, the treatment effective rate is 52.33%. There are no significant differences in age (*P*=0.665, 95%CI 0.47∼3.29, OR=1.24) and gender (*P*=0.151, 95%CI 0.79∼3.90, OR=1.73) except TB treatment effectiveness (*P*=0.007, 95%CI 1.16∼2.84, OR=1.81) between the two groups.

#### 3.1.2 Contribution of different drug regimens from consensus clustering

According to previous studies, different clinical medication regimens affect TB treatment differently. In this study, we try to conduct consensus clustering on the data of 17 drugs in the above 347 samples, select the best drug classification number according to the PAC method based on each medication duration of the samples, and divide each sample into a drug regimen or combination drug regimen group according to the drug use situation. Respectively, compare whether there are significant differences in treatment effectiveness among different drug regimen groups. The influence of age and gender on the difference analysis results is excluded. The results show that the optimal number of drug groups is 3, and 17 drug compounds could be divided into the following three categories, as shown in **Table 3**. Detailed results of the consensus clustering display based on the ConsensusClusterPlus package of R language are shown in **Figure S2**.

**Table 3.**
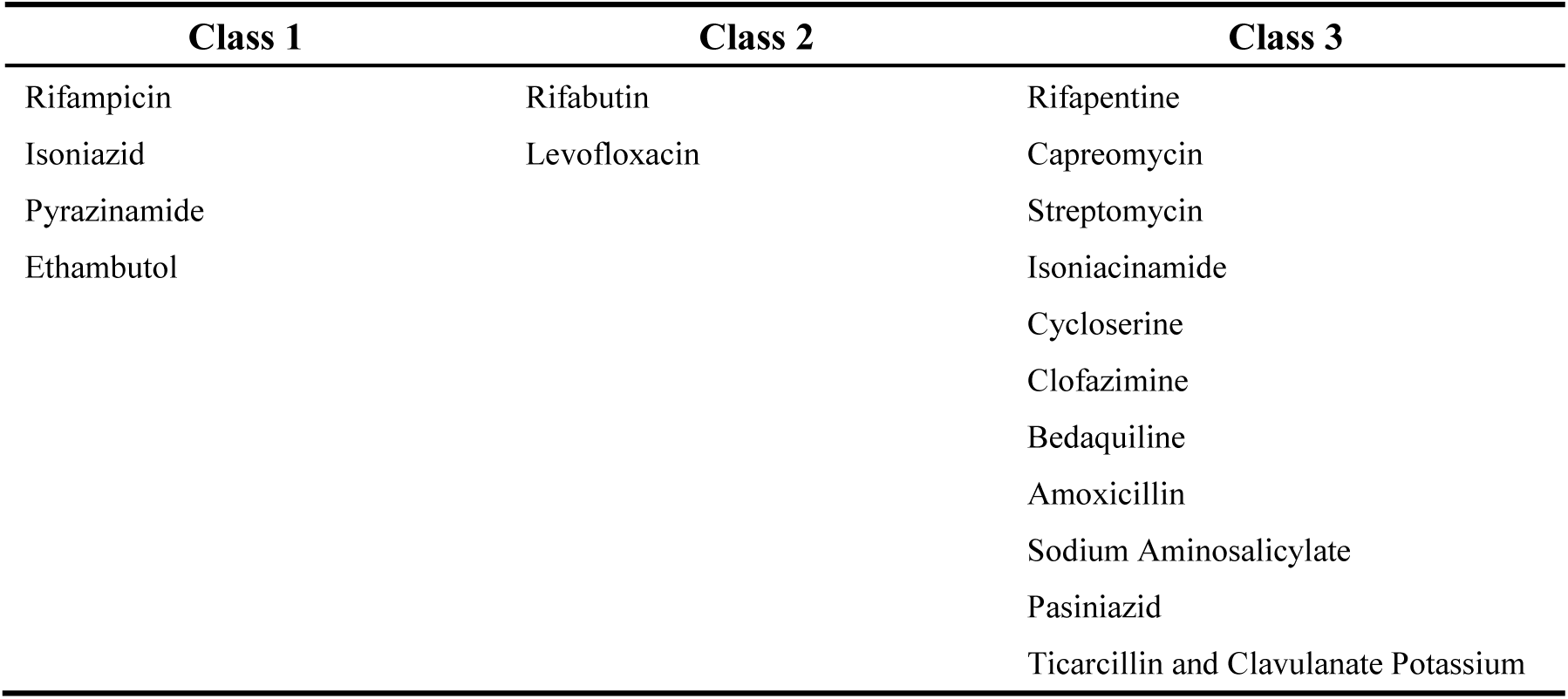
Optimal results of consensus clustering of 17 anti-TB drug compounds.

**Table 4** shows the *P*-value of difference in TB treatment effectiveness among different medication regimens in 347 samples. The results show a significant difference between the “rifabutin and levofloxacin alone or in combination” (class2 group) and “other first- and second-line anti-TB drugs in combination” (class1/3 group) in TB treatment effectiveness (*P*=0.037, 95%CI 0.01∼0.92, OR=0.12). The effective rate of the class2 group is 77.78%, and the class1/3 combined treatment group is 27.78%. There are no significant differences in age (*P*=0.333, 95%CI 0.00∼19.50, OR=0.00) and gender (*P*=1.000, 95%CI 0.01∼Inf, OR=Inf) between the two groups.

**Table 4.** *P*-value of difference analysis in TB therapeutic effectiveness between different medication regimens in seven groups.

|  | class1 | class1/2 | class1/3 | class1/2/3 | class2 | class2/3 | class3 |
| --- | --- | --- | --- | --- | --- | --- | --- |
| class1 |  |  |  |  |  |  |  |
| class1/2 | 0.731 |  |  |  |  |  |  |
| class1/3 | 0.218 | 0.210 |  |  |  |  |  |
| class1/2/3 | 0.512 | 0.734 | 0.219 |  |  |  |  |
| class2 | 0.081 | 0.087 | **0.037** | 0.620 |  |  |  |
| class2/3 | 1.000 | 1.000 | 0.521 | 1.000 | 0.491 |  |  |
| class3 | 1.000 | 1.000 | 1.000 | 1.000 | 0.236 | 1.000 |  |

“/” represents drug combination. For instance, “class1/2” represents class1 and class2 in combination, and all that.

#### 3.1.3 Interaction effect of drug regimens and CD4^+^ T cell counts

In the following work, this study attempts to reveal how the immune level and medication regimen of patients with HIV and TB comorbidities work together to influence the effectiveness of TB treatment. Samples are stratified according to a CD4^+^ T cell count of 42, and Fisher’s exact test is used to analyze further the significance of differences in treatment effect among different drug regimens.

**Table 5** and **Table 6** show the *P*-value of difference in TB treatment effectiveness among different medication regimens within each layer, stratified according to CD4^+^ T cell count of 42. Limited by sample size, when the CD4^+^ T cell count is ≦ 42, the therapeutic effective rate of the group class2 (sample size: 3) is 100%, and that of the group class1/2 (sample size: 91) is 35.2%, the difference of curative effect of the two groups is significant (*P*=0.049, 95%CI 0.00∼1.40, OR=0.00). The two regimens have no significant difference when CD4^+^ T cell count is > 42 (*P*=1.000, 95%CI 0.07∼5.94, OR=0.78). When the CD4^+^ T cell count is ≦ 42, there is no significant difference in efficacy between the class2 group and the class1/3 combined treatment group (*P*=0.143, 95%CI 0.00∼1.88, OR=0.00), neither when the CD4^+^ T cell count is > 42 (*P*=0.319, 95%CI 0.02∼2.52, OR=0.24). There are no significant differences in age and gender between the two groups in the above two stratification, and the analysis results of differences are shown in **Table S2** and **Table S3**. The above conclusions need further confirmation based on increasing the sample size.

**Table 5.** *P*-value of difference analysis in therapeutic effectiveness between different medication regimens in five groups when CD4≦42.

| $CD4 \leq 42$ | class1 | class1/2 | class1/3 | class1/2/3 | class2 |
| --- | --- | --- | --- | --- | --- |
| class1 |  |  |  |  |  |
| class1/2 | 0.626 |  |  |  |  |
| class1/3 | 0.644 | 0.657 |  |  |  |
| class1/2/3 | 1.000 | 1.000 | 1.000 |  |  |
| class2 | 0.071 | **0.049** | 0.143 | 0.400 |  |

“/” represents drug combination. For instance, “class1/2” represents class1 and class2 in combination, and all that.
Statistically significant *P*-values are indicated in bold.

**Table 6.** *P*-value of difference analysis in therapeutic effectiveness between different medication regimens in six groups when CD4>42.

| CD4>42 | class1 | class1/2 | class1/3 | class1/2/3 | class2 | class3 |
| --- | --- | --- | --- | --- | --- | --- |
| class1 |  |  |  |  |  |  |
| class1/2 | 0.129 |  |  |  |  |  |
| class1/3 | 0.370 | 0.066 |  |  |  |  |
| class1/2/3 | 0.707 | 1.000 | 0.356 |  |  |  |
| class2 | 0.427 | 1.000 | 0.319 | 1.000 |  |  |
| class3 | 1.000 | 0.560 | 1.000 | 1.000 | 0.524 |  |

“/” represents drug combination. For instance, “class1/2” represents class1 and class2 in combination, and all that.

### 3.2 Contribution of extensive clinical indicators to the effectiveness of TB treatment in HIV and TB co-infected patients

To comprehensively study the effects of “whether to use a certain drug”, “144 clinical test indicators”, “time interval between two Mycobacterium tuberculosis tests”, and “6 demographic information” on the effectiveness of TB treatment, this study first uses Fisher’s exact test to conduct a single-factor analysis under the precondition of controlling age and gender. Try to screen variables that may significantly impact treatment outcomes and use them for modelling reference in the next step (shown in **3.2.1**). Then, after filtering the “variable-sample” data, three supervised learning classification algorithms, logistic regression (LR), SVM and random forest (RF), are used to construct the two prediction models of TB treatment effectiveness ∼ influencing factors (shown in **3.2.2**).

#### 3.2.1 Single-factor analysis of the effectiveness of TB treatment

The single-factor study shows a significant difference (*P*<0.05) between the effective and ineffective treatment groups in pre-treatment clinical indicators or therapeutic drugs, as shown in **Table 7**. None of the treatment-effective group samples uses Rifapentine, Clofazimine, or Bedaquiline. All samples in this group have normal pre-treatment white blood cell count levels, and none have HBV. In contrast, T-SPOT.TB antigen A (ESAT-6) is abnormal in all samples in the treatment-ineffective group. In conclusion, it is difficult to determine the influence of the six factors above on the effectiveness of TB treatment. Further studies should be conducted based on increasing the sample size. The results of the study on the control variables are shown in **Table S4**.

**Table 7.** Results of difference analysis in pre-treatment clinical indicators and medication regiments between the effective treatment group and the ineffective treatment group in 547 base samples (*P*<0.05)

| Clinical indicators | Treatment-effective group |  | Treatment-ineffective group |  | *P*-value | 95%CI | Odds ratio |
| --- | --- | --- | --- | --- | --- | --- | --- |
|  | Sample number n1 | 1 / (0+1) | Sample number n2 | 1 / (0+1) |  |  |  |
| Cycloserine | 251 | 0.01 | 296 | 0.04 | 0.026 | 1.15~48.72 | 5.25 |
| CD8<sup>+</sup> T cell count | 156 | 0.22 | 193 | 0.33 | 0.042 | 1.01~2.8 | 1.67 |
| Rifabutin | 251 | 0.33 | 296 | 0.25 | 0.029 | 0.44~0.96 | 0.65 |
| Percentage of neutrophils | 245 | 0.53 | 285 | 0.64 | 0.021 | 1.05~2.18 | 1.51 |
| Bicarbonate radical | 43 | 0.67 | 49 | 0.39 | 0.007 | 0.12~0.78 | 0.31 |
| High density lipoprotein cholesterol | 105 | 0.69 | 113 | 0.82 | 0.026 | 1.08~4.26 | 2.12 |
| Age | 251 | 0.11 | 296 | 0.05 | 0.015 | 0.21~0.89 | 0.44 |
| Rifapentine | 251 | **0.00** | 113 | 0.01 | 0.000 | 0.00~0.03 | 0.01 |
| Clofazimine | 251 | **0.00** | 296 | 0.00 | 0.000 | 0.00~0.02 | 0.00 |
| Bedaquiline | 251 | **0.00** | 296 | 0.00 | 0.000 | 0.00~0.02 | 0.00 |
| HBV | 251 | **0.00** | 296 | 0.01 | 0.000 | 0.00~0.03 | 0.01 |
| Leukocyte count | 23 | **0.00** | 26 | 0.08 | 0.000 | 0.01~0.41 | 0.09 |
| T-SPOT.TB Antigen A(ESAT-6) | 77 | 0.99 | 91 | **1.00** | 0.000 | 0.00~0.08 | 0.01 |

The variable “time interval between two *Mycobacterium tuberculosis* tests” is discretized into a binary variable in the univariate analysis, and the classification standard is based on its average value. Less than or equal to the average value is recorded as 0, and greater than the average value is recorded as 1.
“/” represents ratio. For instance, “1 / (0+1)” represents the ratio of the number of samples with binary variables of 1 to those with binary variables of 0 and 1. Therefore, “0.00” in bold represents no sample with the binary variable of 1 in the corresponding group for a specific clinical indicator; “1.00” in bold represents no sample with the binary variable of 0.

Combining the effects of age and gender, the study results show significant differences in age, bicarbonate radical, and HDL cholesterol between effective and ineffective treatment groups. In the samples of patients younger than 60 years old, there are significant differences between treatment-effective and treatment-ineffective groups in the following items, cycloserine is treated or not (*P*=0.027, 95%CI 1.08∼45.90, OR=4.94), rifabutin is treated or not (*P*=0.010, 95%CI 0.40∼0.90, OR=0.60), and pre-treatment CD8^+^ T cell count (*P*=0.012, 95%CI 1.13∼3.37, OR=1.93) and neutrophil percentage (*P*=0.033, 95%CI 1.02∼2.19, OR=1.50). While in samples aged 60 years or older, cycloserine is not used in both the effective and ineffective treatment groups. Rifabutin is used OR not (*P*=0.698, 95%CI 0.26∼9.11, OR=1.58), pre-treatment CD8^+^ T cell count (*P*=1.000, 95%CI 0.05∼6.29, OR=0.66), and neutrophil percentage (*P*=0.751, 95%CI 0.31∼5.77, OR=1.32) show no significant difference between the two groups. For CD8^+^ T cell count, there is a significant difference in male samples younger than 60 years of age (*P*=0.017, 95%CI 1.10∼3.55, OR=1.96) between the treatment response and response groups, not between 60 and older years (*P*=1.000, 95%CI 0.03∼7.36, OR=0.58). There is no significant difference between the female samples under 60 years old (*P*=0.432, 95%CI 0.32∼14.59, OR=2.08) and those over 60 years old (*P*=1.000, 95%CI 0.00∼194.41, OR=0.00).

#### 3.2.2 Multi-factor effectiveness prediction models of TB treatment

At the initial modeling stage, this study tries to construct a prediction model of TB treatment effectiveness using the factors that may affect treatment effectiveness obtained from single-factor analysis as independent variables. However, it is impossible to obtain a well-evaluated model due to the small sample size after filtering or the elimination of independent variables that may have significant contributions to ensure the sample size. Therefore, the two predictive models for TB treatment effectiveness shown below confirm moderate capacity and relatively good efficacy evaluation as far as possible. Still, the selection of independent variables has little correlation with the results of single-factor analysis.

##### Prediction Model I

Screening 547 samples that included demographic information (categorical variable), duration of examination interval, duration of medication, and pre-treatment CD4^+^ T cell count and CD8^+^ T cell count (and the ratio between the two), excluding variables that contain only one category or a total number of classes (or non-zero recorded values) less than five. A matrix containing 347 samples, 18 independent variables, and one dependent variable is obtained. Under five-fold cross-validation, the training set samples corresponding to the test set with the highest prediction accuracy are selected to construct the therapy-effective logistic regression-prediction model I, SVM-prediction model I, and RF-prediction model I. Draw the ROC curves of the training and test sets, respectively, and show the AUC, as shown in Figure 4A. The best AUC values of the test set under the three modeling methods are SVM (0.763), LR (0.659), and RF (0.653) in order from high to low. **Table S5** shows the parameters of the logistic regression prediction model I constructed by stepwise regression, in which the independent variables with significant contributions to prediction include “age” and “CD8^+^ T cell count” (two positive correlations). The importance ranking of variables in RF prediction model I is shown in **Figure S3 A**.

**Figure 4.**
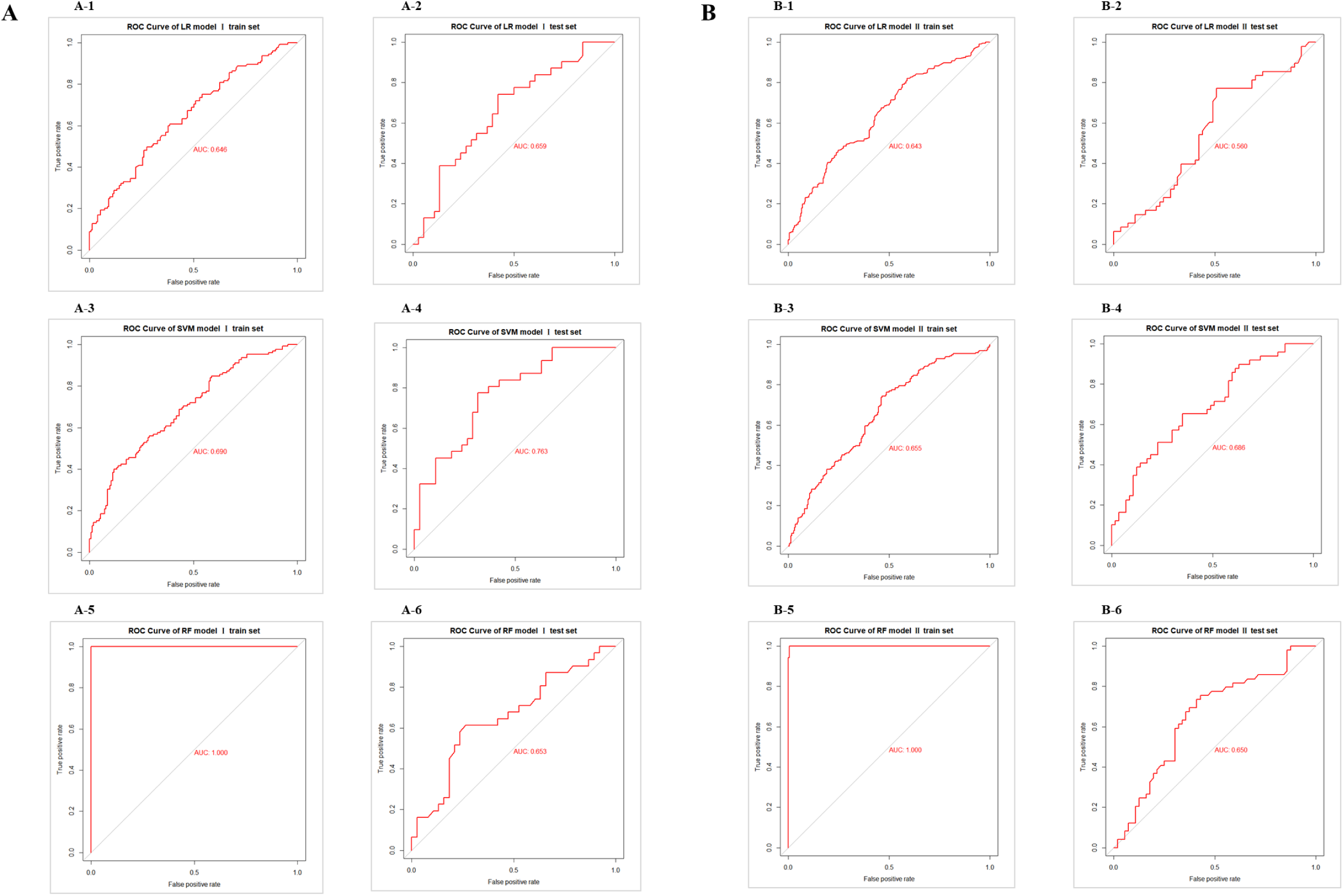
The ROC curve and AUC of the prediction model Ⅰ & Ⅱ based on LR/SVM/RF.

##### Prediction Model II

Among 169 variables, variables with ≥ 500 records are screened, and variables containing only one category or the total number of a category is less than five are excluded. Finally, samples containing missing values of variables are removed from 547 samples, and a matrix containing 527 samples, 31 independent variables, and one dependent variable is obtained. Under five-fold cross-validation, the corresponding training set samples with the highest prediction accuracy of the test set are selected to construct the therapy-effective logistic regression-prediction model II, SVM-prediction model II, and RF-prediction model II. The ROC curves of the training and test sets are plotted respectively, and the AUC is displayed, as shown in Figure 4B. The best AUC values of the test set under the three modeling methods are SVM (0.686), RF (0.650), and LR (0.560) in order from high to low. **Table S6** shows the parameters of logistic regression prediction model II constructed by stepwise regression method, in which independent variables with significant contributions to the prediction include “time interval between two Mycobacterium tuberculosis tests”, “rifabutin”, “sodium aminosalicylate” (three positive correlations) and “cycloserine”, “neutrophilic granulocyte percentage” (two negative correlations). The ranking of the importance of variables in RF prediction model II is shown in **Figure S3 B**.

**Table 8** shows the performance evaluation results of the two prediction models under the three machine learning modeling methods. Figure 5 shows the comparison results of the accuracy of the test set of the prediction model I/II built based on the three algorithms. The results show no significant difference in the accuracy of the two models under any modeling algorithm. In addition, for this batch of research data, the accuracy of the prediction model built based on LR is significantly higher than that built based on SVM. It should not be ignored that the ROC curves of model I /II show serious overfitting in RF modeling. The best AUC of the two models under the three methods is almost less than 0.7, that is, the prediction effect is ordinary.

**Figure 5.**
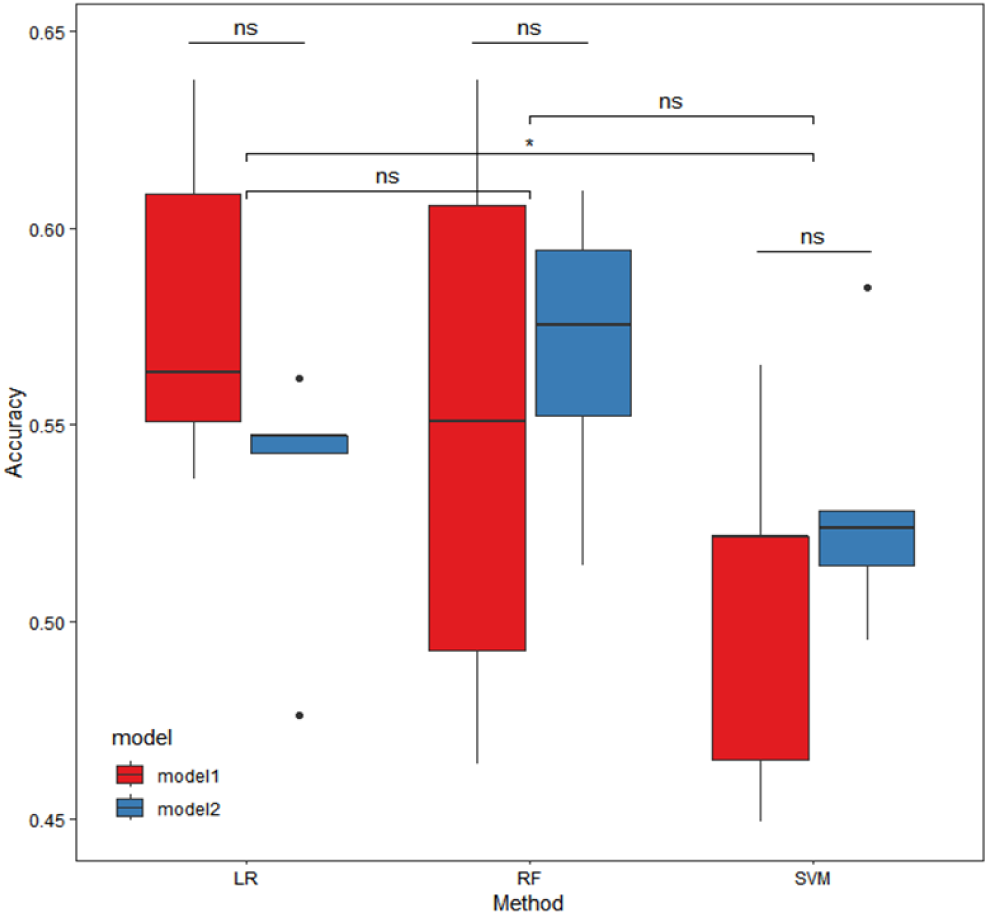
Box plots of accuracy comparison of the test set of prediction model Ⅰ/Ⅱ based on three machine learning algorithms (five-fold cross-validation)

**Table 8.** Results of evaluation indexes of prediction model I/II based on three machine learning algorithms.

| Algorithm | Models | Sets | Sample size | Accuracy | Precision | Recall | F1 score |
| --- | --- | --- | --- | --- | --- | --- | --- |
| Logistic Regression (LR) | prediction | train | 278 | 0.632±0.024 | 0.640±0.031 | 0.410±0.051 | 0.449±0.048 |
|  | model I | test | 69 | 0.579±0.042 | 0.556±0.087 | 0.317±0.054 | 0.442±0.048 |
|  | prediction | train | 422 | 0.634±0.016 | 0.628±0.022 | 0.512±0.011 | 0.564±0.015 |
|  | model II | test | 105 | 0.535±0.034 | 0.500±0.038 | 0.390±0.077 | 0.434±0.053 |
| SVM | prediction | train | 278 | 0.648±0.015 | 0.764±0.023 | 0.319±0.063 | 0.446±0.060 |
|  | model I | test | 69 | 0.505±0.047 | 0.372±0.124 | 0.121±0.059 | 0.176±0.064 |
|  | prediction | train | 422 | 0.620±0.010 | 0.660±0.018 | 0.369±0.043 | 0.472±0.032 |
|  | model II | test | 105 | 0.529±0.034 | 0.484±0.075 | 0.266±0.092 | 0.338±0.080 |
| Random Forest (RF) | prediction | train | 278 | 1.000±0.000 | 1.000±0.000 | 1.000±0.000 | 1.000±0.000 |
|  | model I | test | 69 | 0.550±0.073 | 0.504±0.106 | 0.409±0.093 | 0.449±0.086 |
|  | prediction | train | 422 | 0.994±0.001 | 0.997±0.003 | 0.990±0.004 | 0.993±0.001 |
|  | model II | test | 105 | 0.569±0.037 | 0.543±0.048 | 0.471±0.040 | 0.503±0.031 |

The results of mean ± standard deviation are reported by randomly splitting the training and validation sets for five times.

### 3.3 Differences in clinical indicators changing from abnormal to normal between the effective and ineffective TB treatment groups

Finally, this study analyzes whether there are significant differences in the conversion rate of clinical indicators from abnormal to normal between the effective treatment group and the ineffective treatment group, aiming to find potential or effective therapeutic targets and disease diagnosis sites and provide reference evidence for an in-depth understanding of TB diagnosis and treatment process in patients with AIDS-TB and possible optimization of treatment strategies.

We find only a significant difference in the conversion rate of lymphocyte percentage among all clinical indicators between the effective treatment group and the ineffective treatment group (*P*=0.028, 95%CI 0.32-0.96, OR=0.56). The conversion rate of the effective treatment group is 43.55%, and that of the ineffective treatment group is 30.08%. The analysis results of the top ten clinical indicators with ascending *P*-values are shown in **Table 9**. There is a significant difference in age (*P*=0.018, 95%CI 0.08-0.86, OR=0.29) between the two groups, but no significant difference in gender (*P*=0.353, 95%CI 0.20-1.66, OR=0.60). Further study shows a significant difference in the conversion frequency between the two groups (*P*=0.010, 95%CI 0.27-0.85, OR=0.48) in the samples under 60. The conversion frequency is 45.87% in the treatment-effective group and 28.91% in the treatment-ineffective group. There is no significant difference between the two groups (*P*=0.290, 95%CI 0.31-62.48, OR=3.81) in samples aged 60 years and older.

**Table 9.** Results of difference analysis in clinical indicators from abnormal to normal between effective and ineffective TB treatment groups.

| Clinical Indicators | Treatment-effective group |  | Treatment-ineffective group |  | *P*-value | 95%CI | OR |
| --- | --- | --- | --- | --- | --- | --- | --- |
|  | Sample number | the ratio from | Sample number | the ratio from |  |  |  |
|  | n1 | abnormal to normal | n2 | abnormal to normal |  |  |  |
| Lymphocyte percentage | 124 | 0.44 | 133 | 0.30 | **0.028** | 0.32~0.96 | 0.56 |
| Red blood cell count | 194 | 0.14 | 176 | 0.08 | 0.070 | 0.24~1.05 | 0.51 |
| Prealbumin | 68 | 0.29 | 69 | 0.17 | 0.110 | 0.20~1.22 | 0.51 |
| Smoking | 24 | 0.08 | 31 | 0.26 | 0.159 | 0.65~40.00 | 3.74 |
| Carbamide | 39 | 0.38 | 44 | 0.55 | 0.187 | 0.73~5.07 | 1.90 |
| Hematocrit (HCT) | 206 | 0.07 | 190 | 0.04 | 0.205 | 0.2~1.45 | 0.56 |
| Serum cystatin C | 4 | 0.00 | 9 | 0.44 | 0.228 | 0.30~Inf | Inf |
| Leukocyte count | 98 | 0.41 | 82 | 0.50 | 0.232 | 0.77~2.73 | 1.45 |
| Inorganic phosphorus | 1 | 0.00 | 3 | 1.00 | 0.250 | 0.08~Inf | Inf |
| Globulin | 14 | 0.50 | 17 | 0.71 | 0.288 | 0.44~13.59 | 2.33 |

Statistically significant *P*-values are indicated in bold.

## 4 Discussion

TB is a high incidence of opportunistic chest infection in AIDS patients, and the interaction between the two can accelerate disease progression and eventually cause death. Effective prevention and treatment of AIDS-TB complications is urgent. At present, for the diagnosis, treatment, and prevention of TB in AIDS-TB patients, there is a lack of innovative research methods on the one hand and a lack of high-quality retrospective studies on the efficacy evaluation of treatment programs and the exploration of influencing factors on the other hand. How does the course of AIDS affect the progress of TB treatment, and how is the effectiveness of TB treatment programs affected by the level of the immune system? The real-world research evidence for this is insufficient. Because of the above dilemma, this study focuses on the inpatient data and a small amount of outpatient information on AIDS and TB patients in Shanghai Public Health Clinical Center from 2010 to 2020, using single-factor Fisher’s exact test, hierarchical analysis, and three multi-factor machine learning algorithms. This study comprehensively explores the influencing factors (including drug use and clinical indicators) of TB treatment effectiveness in AIDS-TB co-infected patients and tries to find or confirm TB treatment targets in patients with co-infected patients.

When exploring the influence of immune level and medication regimen on the therapeutic effect of TB in AIDS-TB co-infected patients, the results of a single-factor study show that when the immune level of AIDS-TB co-infected patients is independently judged on the therapeutic effectiveness, the therapeutic effect of TB is limited when the CD4^+^ T cell count is below a certain level, such as 42. Treatment effectiveness is significantly reduced compared to the sample group with CD4^+^ T cell counts greater than 42. When judging the effect of the treatment plan on the treatment effectiveness of AIDS-TB patients alone, if the samples are grouped according to the three-drug clustering results after consistent clustering, the therapeutic effectiveness of the class2 group is significantly higher than that of the class1/3 combined drug group. The possible reason is that, in addition to the limitation of sample size, the efficacy of rifabutin and levofloxacin in the class2 group alone or in combination may be better than that of other first-line and second-line anti-tuberculosis drugs corresponding to class1/3[34–36]. Meanwhile, it cannot be ruled out that the class1/3 combined drug group has more extensive drug resistance, which is more difficult for effective treatment. When considering the influence of immune level and medication plan on treatment effectiveness comprehensively, stratification based on CD4^+^ T cell count being 42 cells/μL and limited by sample size, when CD4≦42, the therapeutic effectiveness of group class2 (sample size is 3) is significantly higher than that of group class1/2 (sample size is 91). There is no significant difference when CD4>42. The possible reasons are that, on the one hand, the reliability of the conclusion needs to be verified in a more expansive and balanced sample range due to the severe imbalance of the sample size. On the other hand, possibly because the class1/2 combination group contains the most effective first-line TB drugs, such as rifampicin and isoniazid, samples with lower immune levels may have been more resistant to such drugs.

However, in each stratification, there is no significant difference between the class2 group and the class1/3 group, which may mean that the uneven distribution of samples causes the difference in conclusions between the population and the stratification levels. In exploring the contribution of clinical indicators to the therapeutic effect of TB in AIDS-TB co-infected patients, single-factor study results show significant differences in age, bicarbonate radical, and high-density lipoprotein cholesterol between effective and ineffective treatment groups. In addition, in samples younger than 60 years of age, there are significant differences in cycloserine use, rifabutin use, pre-treatment CD8^+^ T cell count (male only), and neutrophil percentage between the two groups. These indicators can be used as potential influencing factors for TB treatment effectiveness to construct predictive models. In this study, the model is built under the comprehensive conditions of sufficient samples and good model effect evaluation as far as possible, and the model is interpreted by referring to the results of single factor analysis. The results of the study based on the modeling of three multi-factor machine learning algorithms show that predictive model I uses numerical medication information and immune level to predict the effectiveness of TB treatment and the independent variables that contribute significantly to the effective prediction of TB treatment include “age” and “CD8^+^ T cell count” (two positive correlations). However, the top two independent variables of the importance of contribution are “whether cycloserine is used or not, and age”. Prediction model II uses bifactorial drug use information and multiple clinical indicators to predict TB treatment effectiveness. Among them, independent variables with significant contributions to the effective prediction of TB treatment include “time interval between two Mycobacterium tuberculosis tests”, “rifabutin”, “sodium p-aminosalicylate” (three positive correlations), and “cycloserine” and “neutrophil percentage” (two negative correlations). However, the top two independent variables in the importance of contribution are “whether cycloserine is used or not, and whether sodium para-aminosalicylate is used or not”. The above conclusions are mostly consistent with the results of the single-factor analysis. The results of the model performance evaluation index show that the best AUC of the two models under the three methods is almost less than 0.7. That is, the prediction effect is average. It is worth noting that the accuracy of the prediction model built based on LR is significantly higher than that of the model built based on SVM (both I and II), while the RF modeling has serious overfitting, so logistic regression is relatively the best choice to build the prediction model for this batch of data. The results of the optimal model evaluation in this study are still unsatisfactory, which may indicate some important issues, and the goal of trying to obtain an effective prediction model for TB treatment that is applicable across a wide range of treatment segments without considering the disease course may not be reasonable enough. In addition, the three supervised learning algorithms, including LR with the best modeling performance, may be “too harsh” to interpret the relationship between TB treatment effectiveness and the level of clinical measures, including medication regimen. Moreover, try to use this batch of clinical data to obtain a predictive model with a better evaluation effect, which needs further adjustment.

When analyzing the difference of clinical indicators from abnormal to normal between the TB treatment effective group and the ineffective group in AIDS-TB co-infected patients, the results of the single-factor study show that in the samples under 60 years old, the percentage of lymphocytes in the TB treatment effective group is significantly higher than that in the treatment ineffective group. In comparison, there is no significant difference in the samples over 60 years old. Maybe the ability to recover the immune level is inversely related to age, and the recovery of the immune level may be an insufficient and unnecessary condition for TB treatment to be effective. Lymphocytes are known to produce and carry antibodies to defend against viral infection. The percentage of lymphocytes increases mainly in infectious diseases and decreases mainly in immune deficiency diseases. The analysis results in this study once again demonstrate that lymphocyte percentage could be used as an effective TB therapeutic target in AIDS-TB patients, thus verifying the reliability and validity of the clinical data and experimental design.

The innovation of this retrospective study is to define the interval between every two Mycobacterium tuberculosis test time points of patients as one sample, then define the sample with positive test results at the previous time point and negative test results at the later time point as effective treatment (denoted as 1). The samples with positive test results at the previous time point and positive test results at the later time point are defined as ineffective treatment (denoted as 0), and the factors affecting the effectiveness of treatment are explored in a smaller period based on real data. This way is different from the vast majority of studies that treat a patient as a sample, define the criteria for disease cure in one or several standard courses of treatment, explore the design of trials that affect the effectiveness of treatment, and may uncover helpful information that has been overlooked. In addition, the grouping criteria for therapeutic drugs are based on this batch of clinical data, according to the duration of use of each drug compound, using a consistent clustering method to obtain the combination of drugs prescribed by doctors in real-world conditions, with rare realism. However, some areas for improvement in this study must be addressed. First of all, dividing drug intervals to define samples may ignore the difference in the effect of the same treatment regimen in different stages of disease progression, which is highly likely to cause bias in the study conclusions, and further time series analysis is complex and low feasibility. Secondly, several essential data inclusion criteria need more robust literature or clinical support, such as the minimum number of days of drug use that can be included when screening sample drug use data (defined as seven days according to doctor’s instructions), TB testing criteria (according to doctor’s instructions, one of the microbial tuberculosis test results of any sample type and test type is positive, not only sputum specimen). This deficiency may result in a weak foundation for experimental design. In addition, due to the lack of complete viral load data, it is difficult to accurately measure the HIV course of patients and samples included in this study. Only CD4^+^ T cell counts can be used to assess and stratify patients’ immune system status (or ART treatment level), reducing the richness and reliability of research conclusions. Not only that, the lack of complete drug resistance data prevents conducting further research on drug recommendations for precision therapy. In addition, when pulling the clinical data in this study, the ART treatment data only includes inpatient data, and most outpatient data are missing. It is a pity that the study on the complementary treatment plan of ART and TB treatment could not be carried out. Finally, the interpretability of some clinical routine test indicators regarding how they affect drug effectiveness may require further mechanism exploration, proof, and clinical confirmation. The focus of medical big data research in the real world is how to use the data with uneven structure to obtain the conclusion with as much reference as possible. In the following work, as far as the treatment of AIDS and its complication TB is concerned, prospective experiments can be further designed, continuous, complete, and quantitative AIDS-TB inpatient and outpatient medication data can be collected and recorded, and interactive analysis between AIDS-ART therapy and TB therapy can be carried out. It lays a theoretical foundation for developing and applying personalized treatment in the era of precision medicine.

## 5 Conclusion

TB is the most common cause of AIDS-related death worldwide. Evidence from real-world studies on TB treatment in AIDS-TB co-infected people is insufficient. The retrospective study presented in this paper draws several critical conclusions. Firstly, A CD4^+^ T cell count of 42 cells per μL may serve as an essential and sensitive classification standard for the immune level to assist in evaluating or predicting the effectiveness of TB treatment in comorbidities. Secondly, rifabutin and levofloxacin alone or in combination may be more effective than other first- and second-line anti-TB agents in combination. Thirdly, widespread anti-TB drug resistance may be prevalent in samples with low immune levels (CD4≦42). These samples may be more resistant to the most effective first-line TB drugs, such as rifampicin and isoniazid.

Fourthly, age, bicarbonate radical, high-density lipoprotein cholesterol, pre-treatment CD8^+^ T cell count, neutrophil percentage, rifabutin, and cycloserine may influence the TB treatment effectiveness. Fifthly, the three machine learning modeling methods may be “too strict” to interpret the relationship between clinical test indicators/medication regimen and TB treatment effectiveness. More evidence is needed to support the relationship between clinical indicators before treatment or drug regimens and TB treatment effectiveness. Finally, the percentage of lymphocytes can be used as an effective TB therapeutic target in AIDS-TB patients in combination. The recovery of immune levels may be negatively correlated with age. However, the recovery of the immune level may be an insufficient and unnecessary condition for effective TB treatment. These perspectives may help to supplement the knowledge gaps in relevant clinical aspects and increase the relevant research evidence.

## Author contributions

All authors listed have made a substantial, direct, and intellectual contribution to the work and approved it for publication. Resources: RZ, SL; Investigation: CD, JC, XX, GL; Conceptualization and Methodology: LZ, YL, CD, RZ, JC; Formal analysis, Visualization and Validation: CD; Project administration: LZ, LL, YS; Funding Acquisition: LZ, YS; Supervision: LZ, LL, YS; Writing – original draft: CD, RZ, SL; Writing – review & editing: LZ, LL, YS.

## Data Availability

All data produced in the present study are available upon reasonable request to the authors

## Acknowledgements

We acknowledge the support provided by everyone who contributed to this article. We also acknowledge the support of the Medical Science Data Center in Shanghai Medical College of Fudan University for providing the analysis platform.

## Supplementary materials

**Table S1.** Results of difference analysis between the two groups divided by CD4^+^ T cell counts with stepwise method.

| Grouping criteria<br>(CD4<sup>+</sup> T cell<br>counts) | Sample number n1<br>(CD4 ≤ Grouping<br>criteria) | Sample number n2<br>(CD4 > Grouping<br>criteria) | The ratio<br>of n1 to n2 | *P*-value | 95%CI | Odds<br>ratio |
| --- | --- | --- | --- | --- | --- | --- |
| 40 | 169 | 178 | 0.95 | 0.007 | 1.17~2.89 | 1.84 |
| **42** | **175** | **172** | **1.02** | **0.007** | **1.16~2.84** | **1.81** |
| 10 | 66 | 281 | 0.23 | 0.009 | 1.18~4.05 | 2.15 |
| 41 | 174 | 173 | 1.01 | 0.010 | 1.13~2.78 | 1.77 |
| 13 | 75 | 272 | 0.28 | 0.013 | 1.13~3.60 | 2.00 |
| 39 | 164 | 183 | 0.90 | 0.013 | 1.11~2.73 | 1.73 |
| 11 | 69 | 278 | 0.25 | 0.015 | 1.10~3.65 | 1.98 |
| 43 | 179 | 168 | 1.07 | 0.017 | 1.09~2.68 | 1.71 |
| 12 | 71 | 276 | 0.26 | 0.023 | 1.09~3.53 | 1.94 |
| 37 | 155 | 192 | 0.81 | 0.023 | 1.06~2.62 | 1.66 |
| 19 | 102 | 245 | 0.42 | 0.024 | 1.07~2.93 | 1.76 |
| 15 | 89 | 258 | 0.34 | 0.027 | 1.04~3.01 | 1.76 |
| 17 | 94 | 253 | 0.37 | 0.029 | 1.04~2.96 | 1.75 |
| 50 | 194 | 153 | 1.27 | 0.030 | 1.03~2.55 | 1.62 |
| 51 | 198 | 149 | 1.33 | 0.030 | 1.03~2.53 | 1.61 |
| 47 | 190 | 157 | 1.21 | 0.030 | 1.04~2.56 | 1.63 |
| 38 | 159 | 188 | 0.85 | 0.030 | 1.05~2.58 | 1.64 |
| 14 | 82 | 265 | 0.31 | 0.031 | 1.05~3.14 | 1.80 |
| 44 | 181 | 166 | 1.09 | 0.031 | 1.04~2.55 | 1.62 |
| 45 | 185 | 162 | 1.14 | 0.031 | 1.03~2.53 | 1.61 |
| 8 | 59 | 288 | 0.20 | 0.032 | 1.02~3.66 | 1.91 |
| 2 | 19 | 328 | 0.06 | 0.034 | 1.00~13.65 | 3.23 |
| 9 | 62 | 285 | 0.22 | 0.034 | 1.04~3.62 | 1.91 |
| 28 | 133 | 214 | 0.62 | 0.035 | 1.02~2.60 | 1.63 |
| 29 | 133 | 214 | 0.62 | 0.035 | 1.02~2.60 | 1.63 |
| 123 | 284 | 63 | 4.51 | 0.036 | 1.01~3.30 | 1.82 |
| 36 | 153 | 194 | 0.79 | 0.039 | 1.01~2.51 | 1.59 |
| 49 | 193 | 154 | 1.25 | 0.039 | 1.01~2.49 | 1.58 |
| 46 | 189 | 158 | 1.20 | 0.039 | 1.02~2.51 | 1.60 |
| 18 | 100 | 247 | 0.40 | 0.043 | 1.02~2.80 | 1.68 |
| 128 | 287 | 60 | 4.78 | 0.047 | 0.97~3.24 | 1.77 |
| 30 | 138 | 209 | 0.66 | 0.048 | 0.98~2.47 | 1.56 |
| 53 | 203 | 144 | 1.41 | 0.049 | 0.99~2.45 | 1.56 |
| 54 | 203 | 144 | 1.41 | 0.049 | 0.99~2.45 | 1.56 |
| 52 | 201 | 146 | 1.38 | 0.049 | 1.00~2.46 | 1.56 |
| 124 | 285 | 62 | 4.60 | 0.049 | 0.97~3.19 | 1.75 |
| 34 | 150 | 197 | 0.76 | 0.050 | 1.00~2.48 | 1.57 |
| 35 | 150 | 197 | 0.76 | 0.050 | 1.00~2.48 | 1.57 |

Optimal grouping criteria and related information are indicated in bold.

**Table S2.** Results of difference analysis in control variables between different drug groups based on consensus clustering when CD4≦42.

| **$CD4 \leq 42$** |  |  |  |  |  |  |
| --- | --- | --- | --- | --- | --- | --- |
| **Group of drugs** | **Control variable** | **Sample number n1** | **Sample number n2** | ***P*-value** | **95%CI** | **OR** |
| Group1: class2 | age | 3 | 91 | 0.153 | 0.00~6.77 | 0.10 |
| Group2: class1/2 | gender | 3 | 91 | 1.000 | 0.03~Inf | Inf |
| Group1: class2 | age | 3 | 5 | 0.375 | 0.00~23.40 | 0.00 |
| Group2: class1/3 | gender | 3 | 5 | 1.000 | 0.00~Inf | 0.00 |

**Table S3.** Results of difference analysis in control variables between different drug groups based on consensus clustering when CD4>42.

| **$CD4 > 42$** |  |  |  |  |  |  |
| --- | --- | --- | --- | --- | --- | --- |
| **Group of drugs** | **Control variable** | **Sample number n1** | **Sample number n2** | ***P*-value** | **95%CI** | **OR** |
| Group1: class2 | age | 6 | 64 | 1.000 | 0.07~Inf | Inf |
| Group2: class1/2 | gender | 6 | 64 | 1.000 | 0.16~Inf | Inf |
| Group1: class2 | age | 6 | 13 | 1.000 | 0.00~Inf | 0.00 |
| Group2: class1/3 | gender | 6 | 13 | 1.000 | 0.01~Inf | Inf |

**Table S4.** Results of difference analysis of control variables between the effective treatment group and the ineffective treatment group corresponding to the effective pre-treatment clinical indicators and medication regimens.

| **Clinical indicators** | **Control variable** | ***P*-value** | **95%CI** | **OR** |
| --- | --- | --- | --- | --- |
| **Cycloserine** | age | **0.015** | 0.21~0.89 | 0.44 |
|  | gender | 0.106 | 0.33~1.14 | 0.62 |
| **$CD8^+$ T cell count** | age | **0.008** | 0.09~0.78 | 0.28 |
|  | gender | **0.045** | 0.21~1.01 | 0.47 |
| **Rifabutin** | age | **0.015** | 0.21~0.89 | 0.44 |
|  | gender | 0.106 | 0.33~1.14 | 0.62 |
| **Percentage of neutrophils** | age | **0.023** | 0.22~0.94 | 0.47 |
|  | gender | 0.139 | 0.33~1.16 | 0.62 |
| **Bicarbonate radical** | age | 0.466 | 0.07~2.76 | 0.50 |
|  | gender | 0.662 | 0.05~5.24 | 0.57 |
| **High density lipoprotein cholesterol** | age | 0.202 | 0.06~1.74 | 0.38 |
|  | gender | 0.666 | 0.29~1.95 | 0.76 |
| **Age** | gender | 0.106 | 0.33~1.14 | 0.62 |

Statistically significant *P*-values are indicated in bold.

**Table S5.** Parameters of logistic regression prediction model Ⅰ.

| Coefficients | Estimate | Std.Error | z value | Pr(>\|z\|) |
| --- | --- | --- | --- | --- |
| (Intercept) | -0.7404536 | 0.2247122 | -3.295 | **0.000984\*\*\*** |
| Isoniacinamide | 0.0132609 | 0.0109222 | 1.214 | 0.224700 |
| Cycloserine | -0.2157603 | 0.1151847 | -1.873 | **0.061045.** |
| Sodium aminosalicylate | 0.1472285 | 0.1133682 | 1.299 | 0.194055 |
| Age | 1.5180879 | 0.6780730 | 2.239 | **0.025167\*** |
| Gender | 0.6707818 | 0.4081069 | 1.644 | 0.100250 |
| CD8<sup>+</sup> T cell count | 0.0008933 | 0.0004068 | 2.196 | **0.028086\*** |

Significance: 0 '\*\*\*' 0.001 '\*\*' 0.01 '\*' 0.05 '.' 0.1 ' ' 1

**Table S6.** Parameters of logistic regression prediction model II.

| Coefficients | Estimate | Std.Error | z value | Pr(>\|z\|) |
| --- | --- | --- | --- | --- |
| (Intercept) | 0.221921 | 0.348846 | 0.636 | 0.5247 |
| Time interval for TB detection | 0.003507 | 0.001734 | 2.022 | **0.0431\*** |
| Rifabutin | 0.583400 | 0.222029 | 2.628 | **0.0086\*\*** |
| Ethambutol | -0.525134 | 0.336162 | -1.562 | 0.1183 |
| Cycloserine | -3.103263 | 1.265058 | -2.453 | **0.0142\*** |
| Sodium aminosalicylate | 2.160373 | 1.099459 | 1.965 | **0.0494\*** |
| neutrophilic granulocyte percentage | -0.436272 | 0.204830 | -2.130 | **0.0332\*** |

Significance: 0 '\*\*\*' 0.001 '\*\*' 0.01 '\*' 0.05 '.' 0.1 ' ' 1

**Figure S1.**
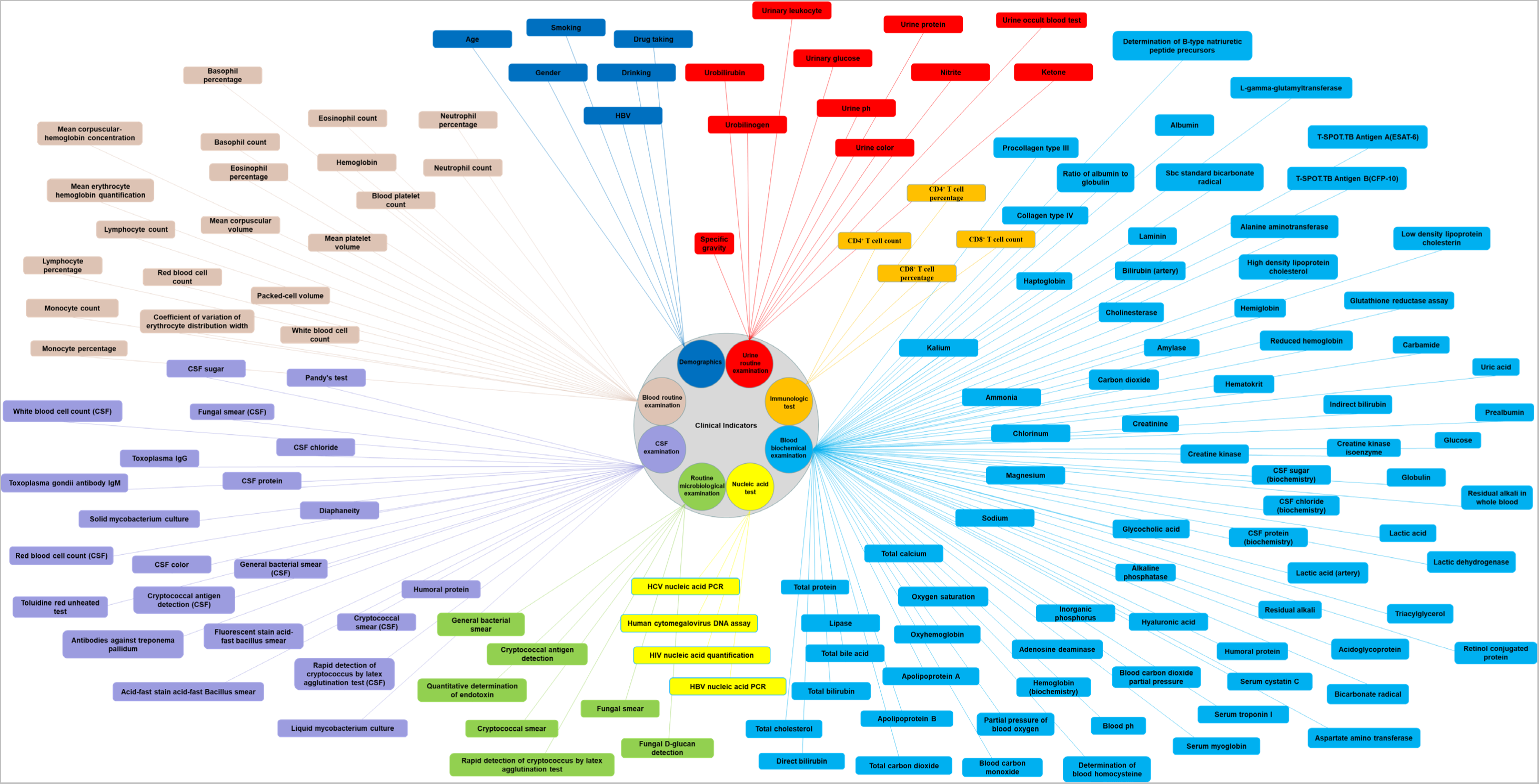
150 clinical features from 8 categories (include demographics)

**Figure S2.**
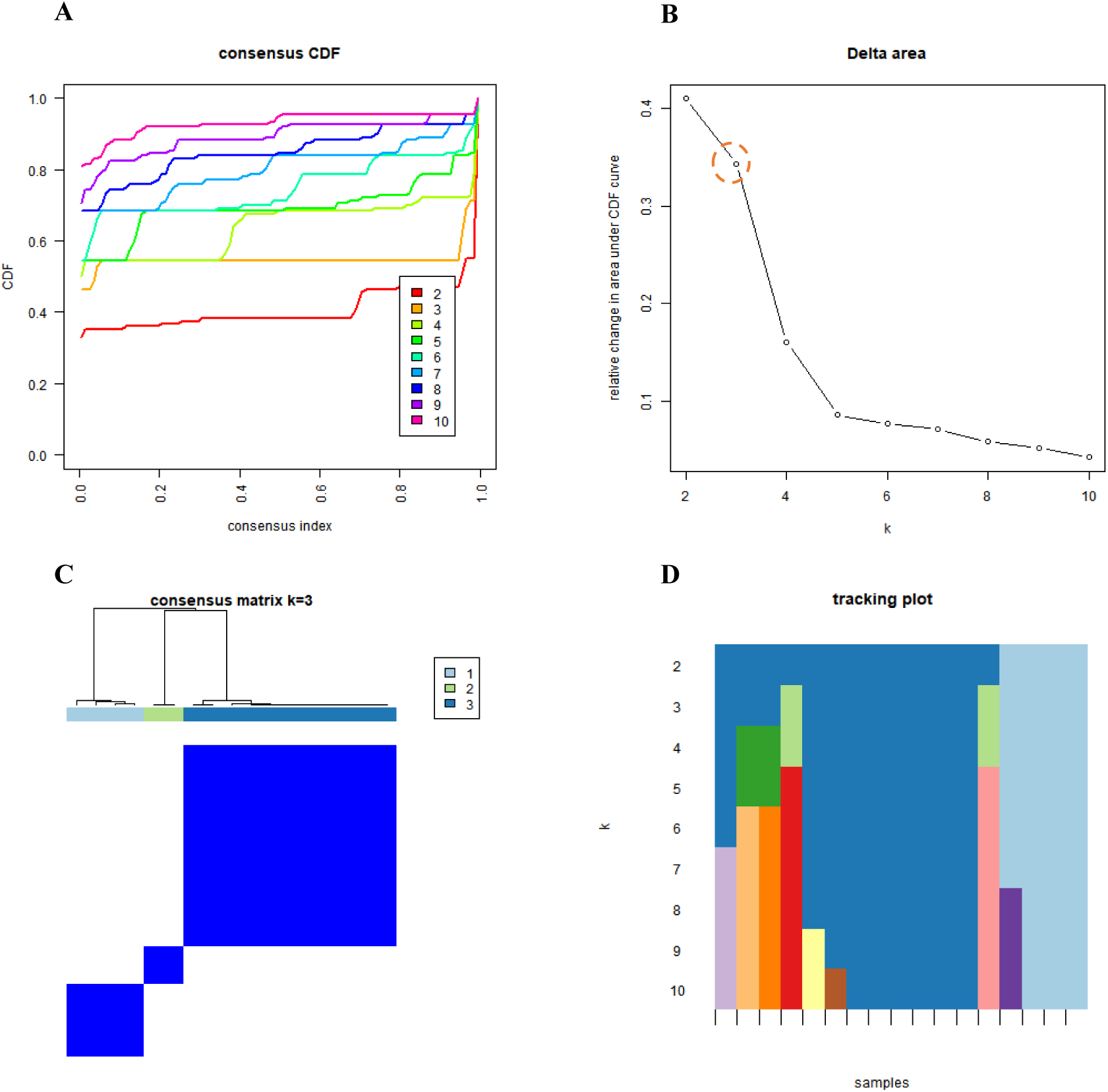
Results of consensus clustering display based on ConsensusClusterPlus package of R.

Figure S2 shows the results generated by consistent clustering of 17 pharmaceutical chemical components of all samples using the R language ConsensusClusterPlus package. **FIG. S2A** shows the cumulative distribution function (CDF) graph. The cluster analysis results are more reliable when the CDF gradient is smaller and corresponds to the k value. **FIG. S2B** represents the delta area diagram, which shows the relative change of the area under the CDF curve compared with *k* and *K*-1. In general, the point (inflection point) that starts to slow down, *k*=5, can be used as the more suitable cluster number. However, according to the clustering performance shown in the heat map of this batch of data, *k*=3 is finally selected as the best group number by the more reliable PAC method. **FIG. S2C** shows the matrix heat map when *k*=3, the rows and columns of the matrix are samples, and the values of the consistency matrix are represented by white to dark blue from 0 (impossible to cluster together) to 1 (always clustered together), with good clustering effect and almost no noise. The optimal *k* value obtained by combining the PAC method is 3. The black stripes at the bottom of **FIG. S2D** represents samples, showing the classification of samples belonging to different values of *k*. Different color blocks represent different categories, and the classification is more stable when *k*=3.

**Figure S3.**
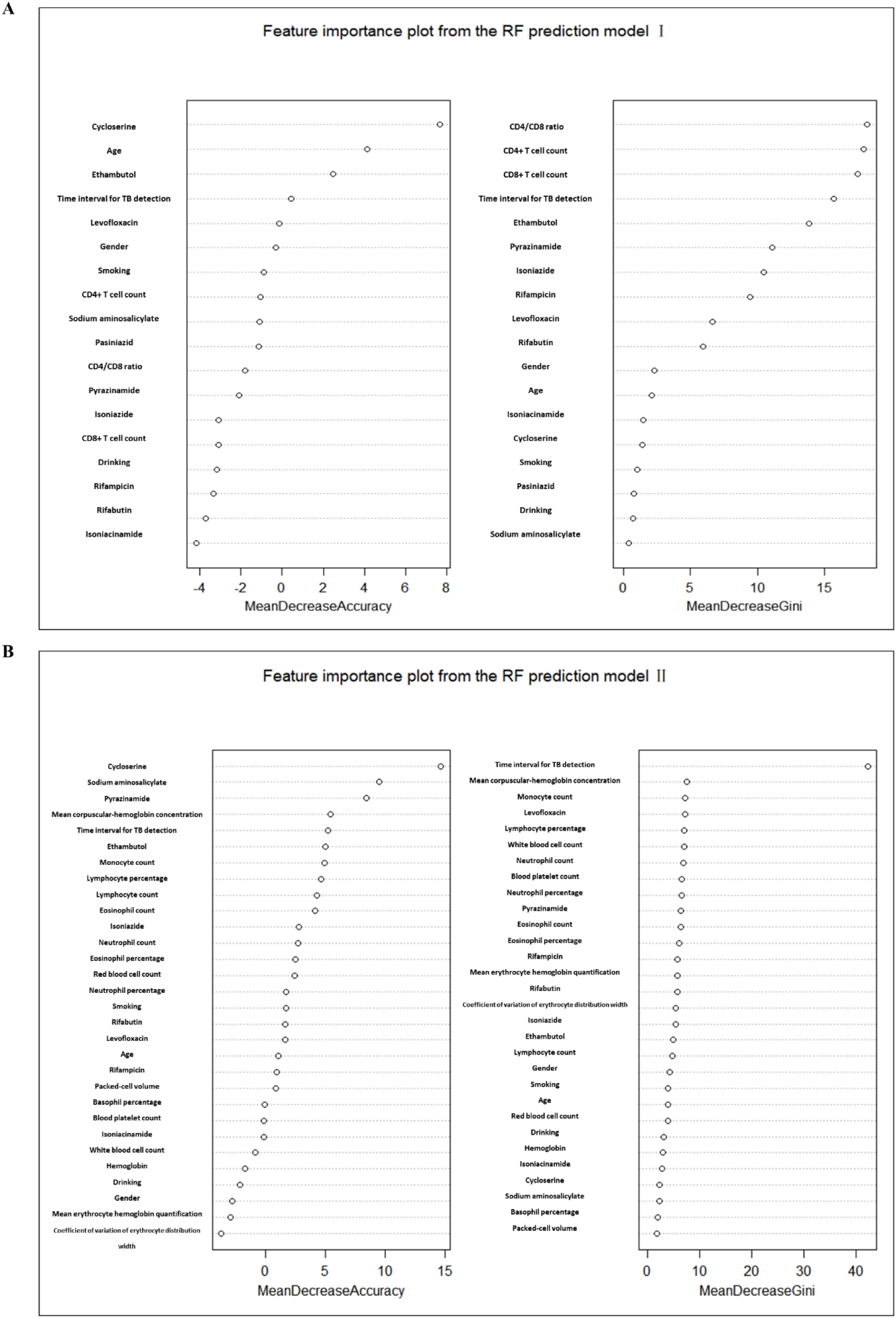
Feature importance plots from the RF prediction model Ⅰ & Ⅱ.

